# GX-I7(rhIL-7-hyFc, efineptakin alfa), a long-acting IL-7, safely and effectively increased peripheral CD8^+^ and CD4^+^ T cells and TILs in patients with solid tumors

**DOI:** 10.1101/2024.02.12.23299638

**Authors:** Gun Min Kim, Sojeong Kim, Myung Ah Lee, Mi-Sun Byun, Donghoon Choi, Se Hwan Yang, Jung Won Woo, Young Chul Sung, Eui-Cheol Shin, Su-Hyung Park, Tae Won Kim, Joohyuk Sohn

## Abstract

**Purpose:** GX-I7 (rhIL-7-hyFc, efineptakin alfa) is a hybrid Fc-fused long-acting recombinant human interleukin-7 (IL-7) developed by Genexine, Inc. with the aim of correcting T-cell deficiency, thereby strengthening the immune response to fight against cancer. This Phase 1b trial (NCT03478995) was designed to assess the safety, tolerability, pharmacokinetics (PK), and pharmacodynamics (PD) of GX-I7 in patients with locally advanced or metastatic solid tumors.

**Methods:** This study consisted of two phases: dose-escalation and expansion. Eight dose groups were administered GX-I7 intramuscularly at doses ranging from 60 to 1700 μg/kg every three or six weeks.

**Results:** All regimens were safe and well tolerated, with the most frequently reported adverse drug reactions being injection site reactions, which were manageable with or without pharmacological intervention. GX-I7 demonstrated dose-dependent increases in the maximum serum concentration (C_max_) and area under the curve up to the last measurable concentration (AUC_last_). In addition, a dose-dependent increase in circulating CD8^+^/CD4^+^ T cells was observed. In five patients who consented for biopsy, a statistically significant increase in tumor-infiltrating CD8^+^/CD4^+^ T cell lymphocytes after GX-I7 treatment was observed.

**Conclusion:** These findings support the use of GX-I7 as a safe and effective T cell-amplifying agent capable of correcting T cell deficiencies. GX-I7 is expected to result in better clinical outcomes when used in combination with other anti-cancer agents by creating a better environment for immune checkpoint inhibitors and anti-cancer treatments to fight cancer.

## INTRODUCTION

Surgery and chemotherapy have long been the two major modalities for cancer treatment, while chemotherapy is the sole option for patients with non-resectable advanced or metastatic solid tumors. However, with the emergence of immune therapies, such as immune checkpoint inhibitors (ICI), T cells have become a central focus in engaging the immune system in battle against cancer(1). Previous studies have revealed that lymphopenia (2), including anti-cancer treatment-induced lymphopenia, is associated with inferior survival outcomes (3-5). Furthermore, a low response to ICI therapy has been observed in patients with low lymphocyte count (6). As T cells play a central role in the immune response to cancer, correcting lymphopenia and increasing tumor infiltrating lymphocytes (TILs) are crucial in enhancing the immune response to cancer, thereby improving treatment outcomes (7, 8).

GX-I7 (rhIL-7-hyFc, efineptakin alfa) is a hybrid Fc-fused long-acting recombinant human interleukin-7 (IL-7) developed by Genexine, Inc., with the aim of correcting T cell deficiency, thereby strengthening the immune response to fight cancer and creating a favorable environment for anti-cancer treatment or ICIs. IL-7 is a homeostatic cytokine that plays an important role in the development, maintenance, proliferation, and regeneration of T-cells (9, 10). Despite the clinical benefits of IL-7 in various therapeutic areas, including cancer, the development of IL-7 as a commercial drug is limited by its short half-life (11-19).

GX-I7 effectively lengthens the IL-7 half-life by six times compared to that of rhIL-7 via neonatal Fc receptor (FcRn)-mediated recycling (20, 21). In vivo, GX-I7 led to an increase in peripheral lymphocyte counts and TILs when administered either in monotherapy or combination therapy with cyclophosphamide (CPA) or ICI (e.g., anti-PD-(L)1 or anti-CTLA-4 antibody), and showed anti-tumor efficacy in murine syngeneic tumor models (22).

With in vivo and clinical Phase 1 study results supporting further development of GX-I7 to correct T cell deficiencies, this study was designed to assess safety, tolerability, pharmacokinetics (PK) and pharmacodynamics (PD) of GX-I7 in patients with locally advanced or metastatic solid tumors.

## RESEARCH DESIGN AND METHODS

### Ethics statement

This study was approved by the institutional review board of each research site (Yonsei University Health System Severance Hospital, Seoul Asan Medical Center and Catholic Medical Center) and conducted in full accordance with the Declaration of Helsinki and Korean Good Clinical Practice (www.clinicaltrials.gov registration number: NCT03478995). Written informed consent was obtained from all patients prior to conducting any study-related procedures.

### Study design

This was a Phase 1b, open-label, dose-escalation study to evaluate the safety, tolerability, PK, and PD of GX-I7 in patients with locally advanced or metastatic solid tumors. The primary objectives were to determine the recommended phase 2 dose (RP2D) and maximum tolerated dose (MTD) and to characterize dose-limiting toxicities (DLTs). Classic 3+3 design was adopted for sequential dose escalation, and the DLT evaluation period was 3 weeks.

### Patients

Eligible patients were aged ≥ 19 years and had an Eastern Cooperative Oncology Group (ECOG) performance status of 0 or 1, an estimated life expectancy of > 12 weeks, and appropriate hematologic, hepatic, and end-organ function values. Patients had histological documentation of locally advanced, recurrent, or metastatic incurable solid tumors that had failed to know or available therapy, or for whom standard therapy was considered inappropriate. Exclusion criteria included any anti-tumor therapy within three weeks and previous use of an ICI or immunomodulatory monoclonal antibody (mAb)- derived drug within 12 weeks prior to the initiation of study treatment.

### Treatment

Patients were enrolled to receive GX-I7 intramuscularly at three- or six-week intervals. The dose was escalated, and a total of eight dose levels were tested, each with at least three patients, at doses of 60, 120, 240, 480, 720, 960, 1200, and 1700 μg/kg at three-week intervals. Out of the eight tested dose levels, two (720 and 1200 μg/kg) were selected as potential RP2D for expansion, and up to six additional patients were enrolled at these doses for the expansion phase. At a dose expansion of 1200 μg/kg, the efficacy of GX-I7 was evaluated at both three- and six-week intervals. For biopsy-consented patients, tumor tissue was collected both pre and three–five weeks after post-GX-I7 administration to test changes in TILs.

### Safety, tolerability, and immunogenicity assessments

The safety and tolerability of GX-I7 were assessed throughout the study by adverse event (AE) monitoring using the NCI Common Terminology Criteria for Adverse Events (CTCAE) (version 4.03). Undesirable symptoms, clinically significant abnormalities in clinical laboratory tests, vital signs, physical examination, or DLT, as well as the emergence of antibodies (anti-drug antibody [ADA] and neutralizing antibody [nAb]) were included. Treatment-emergent adverse events (TEAEs) were analyzed. The presence of antibodies against GX-I7 was determined using a validated enzyme-linked immunosorbent assay (ELISA), as described in the GX-I7 Phase 1 First-in-human (FIH) study (22). All patients who tested positive for ADA were analyzed for its neutralizing property (nAb).

### Efficacy

Solid tumor assessments were performed by CT scan or MRI every 2 cycles up to cycle 8, and every four cycles or as clinically indicated thereafter. Tumor response and progression were evaluated according to the Response Evaluation Criteria in Solid Tumors (RECIST) (version 1.1).

### PK analysis

Blood samples for PK analysis were collected during Cycle 1 while the patients were hospitalized at various time points, including pre-dose and 0.5, 6, 12, 24, 48, and 72hours post-dose. Additional blood collections were performed on Cycle 1 Day 8 (C1D8), C1D15, and C2D1 for patients administered GX-I7 every 3 weeks and C1D8, C1D15, C1D22, and C2D1 for patients administered GX-I7 every 6 weeks. Serum IL-7 concentrations were measured using the Quantikine HS ELISA Human IL-7 immunoassay kit (R&D Systems, Minneapolis, MN), and the detailed analysis method was based on a previously reported phase 1 study (22). The PK parameters were derived by a non-compartmental method using Phoenix WinNonlin software (version 7.0, Certara, St. Louis, MO, USA). In addition, the correlation between the administered dose and AUC_last_ and C_max_ was analyzed.

### PD analysis

The PD variables were changes in absolute lymphocyte counts (ALC) and subsets of immune cells (T, B, and NK cells) in the blood assessed on a weekly basis at pre-dose, C1D1, and C2D1. Thereafter, it was performed on day 1 (pre-dose) of every cycle and at the treatment discontinuation (DC) visit. Absolute T, B, and NK cell counts were calculated using ALCs multiplied by the proportions of CD3^+^, CD19^+^ and CD56^+^ cells against total lymphocytes, and absolute CD4^+^ and CD8^+^ T cells were calculated using ALCs multiplied by the proportions of CD4^+^ and CD8^+^ T cells against total CD3^+^ T cells, respectively. Ki67^+^ and CD127^+^ T cell counts were calculated by multiplying the proportion of Ki67^+^- or CD127^+^-expressing cells by total CD4^+^ and CD8^+^ T cells, respectively. All analyses were performed using multicolor flow cytometry. The following anti-human fluorochrome-conjugated antibodies were used: anti-CD3-Alexa Fluor 700 (UCHT1), anti-IL-7Rα-BV421 (A019D5), anti-CD4-PE-Cy5 (OKT4), anti-CD45RA-APC-Cy7 (HI100), anti-CCR7-BV785 (G043H7), anti-CD25-BV650 (BD96; BioLegend), anti-CD19-BV711 (SJ25C1), anti-CD14-BV711 (MφP9), anti-CD8-Alexa Fluor 700 (RPA-T8; all from BD Biosciences), and anti-CD56-APC (CMSSB; Thermo Fisher). Simultaneously, dead cells were stained using a Live/Dead Fixable Red Dead Cell Stain Kit (Thermo Fisher Scientific). The cells were fixed and permeabilized using a Foxp3 Staining Buffer Set (Thermo Fisher), and intracellular molecules were stained (4 °C, 20 min) using anti-Ki-67-PE-Cy7 (20Raj1) and anti-Foxp3-PE (236A/E7; Thermo Fisher) (Supplementary material). To calculate the maximum ALC and the area under the ALC-time curve until the last sampling time, the Phoenix WinNonlin software was used after correcting the baseline values. The change in TILs at pre and post GX-I7 administration was evaluated in patients who consented to biopsy using Multiplex IHC analysis (Supplementary Material).

### Statistical analysis

All demographic characteristics and PK and PD parameters were summarized using descriptive statistics. Continuous variables were summarized using the number of observations, mean, standard deviation, coefficient of variation, median, and range as appropriate. Categorical values were summarized using the number of observations and percentages as appropriate. Data were summarized for the dose cohort, expansion cohort, and total patient population. All assessments before the first dose of the study drug were considered as baseline. If multiple baseline assessments were performed, the most recent assessment was used for statistical analysis. A parametric or nonparametric method was used based on the results of the normality test. All analyses were 2-sided, with a significance level of 5%, and a 95% confidence interval (CI) was calculated. All statistical analyses were performed using SAS® Version 9.4 (SAS Institute, Cary, NC, USA).

## RESULTS

### Patients disposition and baseline characteristics

Out of 42 patients screened across three hospitals, 35 patients were enrolled in the study and received at least one dose of GX-I7, with seven patients being excluded because they either did not satisfy the eligibility criteria (five patients) or withdrew their consent (two patients). In the dose escalation phase, according to the classical 3 + 3 design, 3 patients were assigned to each group. In the 1700 μg/kg group, only two patients were enrolled because of the occurrence of one DLT. The group receiving 720 μg/kg, which showed the highest increase in ALC fold change, and 1200 μg/kg as the maximum tolerated dose (MTD) was selected as the potential recommended phase 2 dose (RP2D). Three patients receiving 720 μg/kg group every 3 weeks (q3w), three patients receiving 1200 μg/kg q3w, and six patients receiving 1200 μg/kg q6w were enrolled in the dose expansion (Figure 1). A dose expansion group receiving GX-I7 q6w was included after determining that since the ALC remained elevated at the time of the q3w repeated dosing, it was not expected that frequent administration of GX-I7 would bring additional benefits. The median age (range) was 58 (40-75) years; 54% were male, and all were Asian. Baseline ECOG performance status was 0 in 49% of patients, and the most common site of the primary tumor was colon/rectal in 66% of patients, followed by the breast in 11%, and the ovary in 9% of patients. At the time of study enrollment, metastatic lesions were commonly found in the lungs (32 patients), followed by the liver (24 patients), and lymph nodes (14 patients). All subjects enrolled in the study were at stage IV with a third or more lines of treatment. In previous therapies, the majority of subjects had been treated with antimetabolite, targeted, and platinum-based agents. The subjects discontinued their previous treatment primarily because of disease progression (PD). The demographic and baseline characteristics are summarized in Table 1.

**Table 1.** Baseline characteristics.

| Group | GX-I7<br>60 µg/kg<br>(n=3) | GX-I7<br>120 µg/kg<br>(n=3) | GX-I7<br>240 µg/kg<br>(n=3) | GX-I7<br>480 µg/kg<br>(n=3) | GX-I7<br>720 µg/kg<br>(n=6) | GX-I7<br>960 µg/kg<br>(n=3) | GX-I7<br>1200 µg/kg<br>(n=12) | GX-I7<br>1700 µg/kg<br>(n=2) | Total<br>(n=35) |
| --- | --- | --- | --- | --- | --- | --- | --- | --- | --- |
| **Age (years)** |  |  |  |  |  |  |  |  |  |
| Median (Min, Max) | 58 (52, 64) | 58 (55, 58) | 59 (52, 75) | 44 (44, 48) | 57 (45, 71) | 58 (45, 65) | 62 (43, 75) | 54 (48, 60) | 58 (40, 75) |
| **Gender, N (%)** |  |  |  |  |  |  |  |  |  |
| Male | 2 (67) | 0 | 3 (100) | 2 (67) | 4 (67) | 1 (33) | 6 (50) | 1 (50) | 19 (54) |
| **Race, N (%)** |  |  |  |  |  |  |  |  |  |
| Asian | 3 (100) | 3 (100) | 3 (100) | 3 (100) | 6 (100) | 3 (100) | 12 (100) | 2 (100) | 35 (100) |
| **ECOG performance status, N (%)** |  |  |  |  |  |  |  |  |  |
| 0 | 1 (33) | 1 (33) | 1 (33) | 0 | 5 (83) | 2 (67) | 5 (42) | 2 (100) | 17 (49) |
| 1 | 2 (67) | 2 (67) | 2 (67) | 3 (100) | 1 (17) | 1 (33) | 7 (58) | 0 | 18 (51) |
| **Site of Primary Tumor, N (%)** |  |  |  |  |  |  |  |  |  |
| Breast | 1 (33) | 0 | 0 | 1 (33) | 0 | 0 | 2 (17) | 0 | 4 (11) |
| Colon/Rectal | 1 (33) | 2 (67) | 2 (67) | 2 (67) | 4 (67) | 3 (100) | 7 (58) | 2 (100) | 23 (66) |
| Ovary | 0 | 1 (33) | 0 | 0 | 0 | 0 | 2 (17) | 0 | 3 (9) |
| Other | 1 (33) | 0 | 1 (33) | 0 | 2 (33) | 0 | 1 (8) | 0 | 5 (14) |
| **Location of Metastatic, N** |  |  |  |  |  |  |  |  |  |
| Abdominal Wall | 0 | 1 | 0 | 0 | 1 | 1 | 1 | 0 | 4 |
| Adrenal Gland | 1 | 0 | 1 | 0 | 0 | 0 | 1 | 0 | 3 |
| Bone | 1 | 0 | 0 | 1 | 1 | 0 | 1 | 0 | 4 |
| Brain | 0 | 0 | 0 | 1 | 0 | 0 | 1 | 0 | 2 |
| Liver | 2 | 2 | 2 | 2 | 1 | 3 | 10 | 2 | 24 |
| Lung | 3 | 2 | 3 | 3 | 5 | 3 | 11 | 2 | 32 |
| Lymph Node | 2 | 2 | 1 | 2 | 3 | 2 | 2 | 0 | 14 |
| Pleura | 1 | 0 | 0 | 0 | 0 | 0 | 1 | 0 | 2 |
| Other | 2 | 0 | 2 | 1 | 2 | 2 | 4 | 0 | 13 |

**Figure 1.**
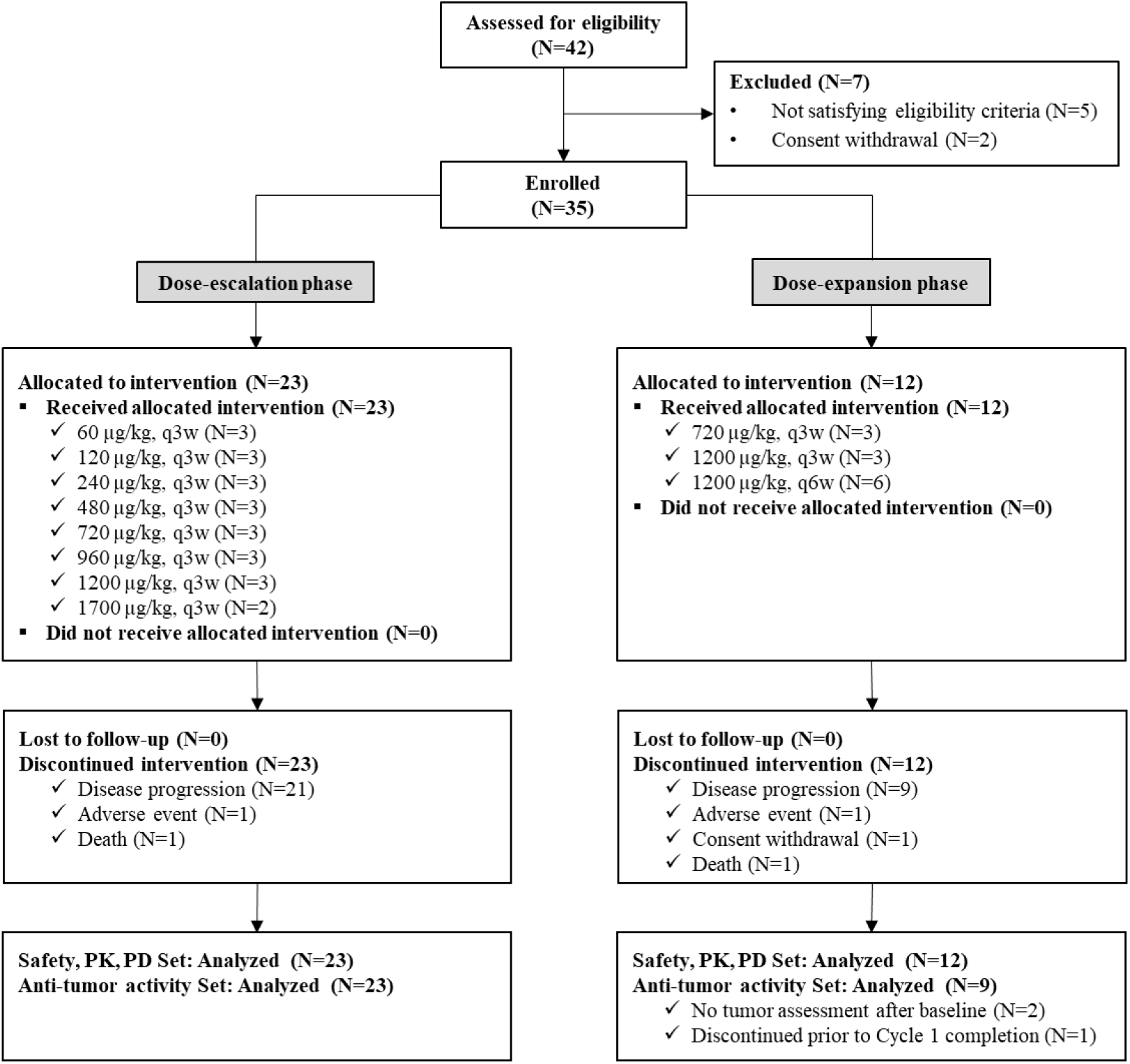
Patient disposition. Patients in the dose-escalation stage were treated with 60, 120, 240, 480, 720, 960, 1200 and 1700 μg/kg of GX-I7 intramuscularly every 3 weeks (Q3W). In the dose-expansion stage, patients received a 720 and 1200 μg/kg dose of GX-I7 intramuscularly every 3 weeks (Q3W) or every 6 weeks (Q6W), which were selected in the dose-escalation stage.

### Safety

The safety analysis set included patients who received at least one dose of GX-I7 during the clinical trial. Safety data from all 35 enrolled patients were used to assess the safety profiles. GX-I7 intramuscular administration was safe and well tolerated across doses ranging from 60 to 1200 μg/kg, except for the 1700 μg/kg dose. A total of 225 TEAEs occurred in all the 35 patients (100%). Among these, 116 adverse drug reactions (ADRs) occurred in 28 of 35 patients (80.0%), with the majority rated as Grade 1 and 2 (68 and 43 cases, respectively). The most frequently reported ADR was injection site reaction (61 cases in 25 patients (71.4%)), followed by pyrexia with 21 cases in 12 patients (34.3%), then rash with 8 cases in 7 patients (20.0%). All 61 reported injection site reactions were classified as grade 1 or 2, and most were spontaneously resolved, with 32 cases being resolved with medications. No cases of grade 3 or 4 injection site reactions were observed.

In the 720, 1200, and 1700 μg/kg groups, grade 3 or higher ADRs were reported: fatigue (720 and 1200 μg/kg), hypersensitivity, and anaphylactic reactions (1700 μg/kg). Due to grade 3 fatigue and anaphylactic reaction, two patients discontinued GX-I7 permanently. No GX-I7 related death have been reported.

The dose-limiting toxicity of GX-I7 was assessed in 35 patients for all dose groups, and at 1700 μg/kg, one out of two patients experienced DLT, which was a case of Grade 3 or higher hypersensitivity. Therefore, the MTD was determined to be 1200 μg/kg, and this dose administered every 6 weeks was selected as RP2D. Regardless of the administration interval, the incidence of ADA increased on day 21, and no ADA-related AE were observed. A summary of the safety profile is presented in Table 2.

**Table 2.**
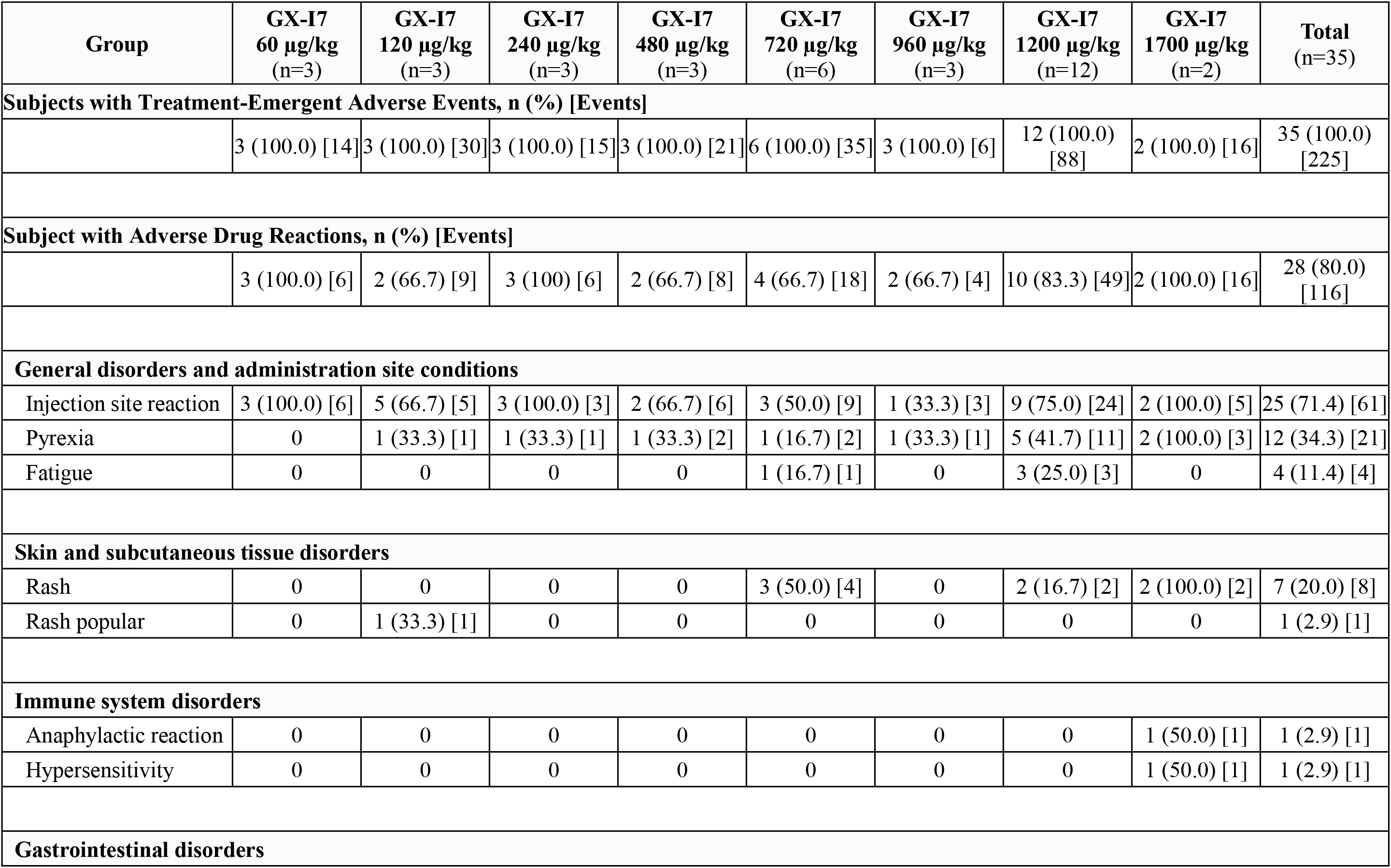
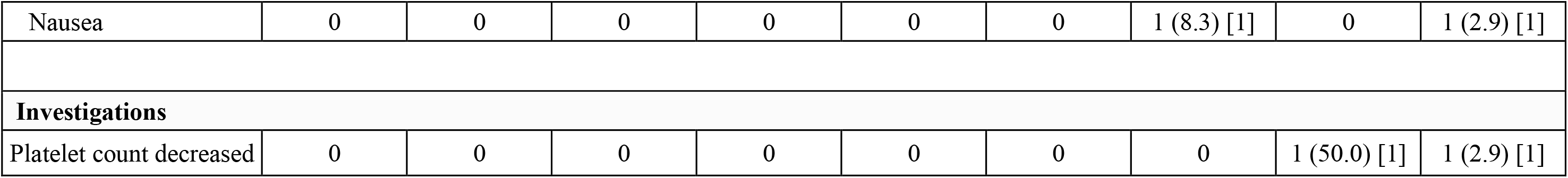
Summary of safety profiles.

### Tumor response

Of the 35 patients with solid tumors who underwent dose escalation and expansion, 6 patients (18.75%) showed disease control as stable disease (SD). Among these six patients, the doses received were 60, 120, 240, 720, and 1200 μg/kg, as shown in Supplementary Table S1. The median PFS was 5.69 weeks (95% CI; 5.13, 5.98).

### Pharmacokinetics (PK)

This study conducted a PK analysis of 35 patients from all cohorts. As shown in Figure 2A, serum concentration was measured at various time points after intramuscular administration of GX-I7, ranging from 60 to 1700 μg/kg. The PK profile of GX-I7 showed a dose-dependent response. The peak serum concentration (C_max_) was reached at 11–47.5 hours post dosing, with peak serum concentration ranging from 2.11 to 76.5 ng/mL, and a half-life lasting from 60.8 to 139.7 hours (Supplementary Table S2). AUC_last_ and C_max_ increased linearly with dose (R^2^ = 0.9174 and 0.9657, respectively; Figure 2B and 2C).

**Figure 2.**
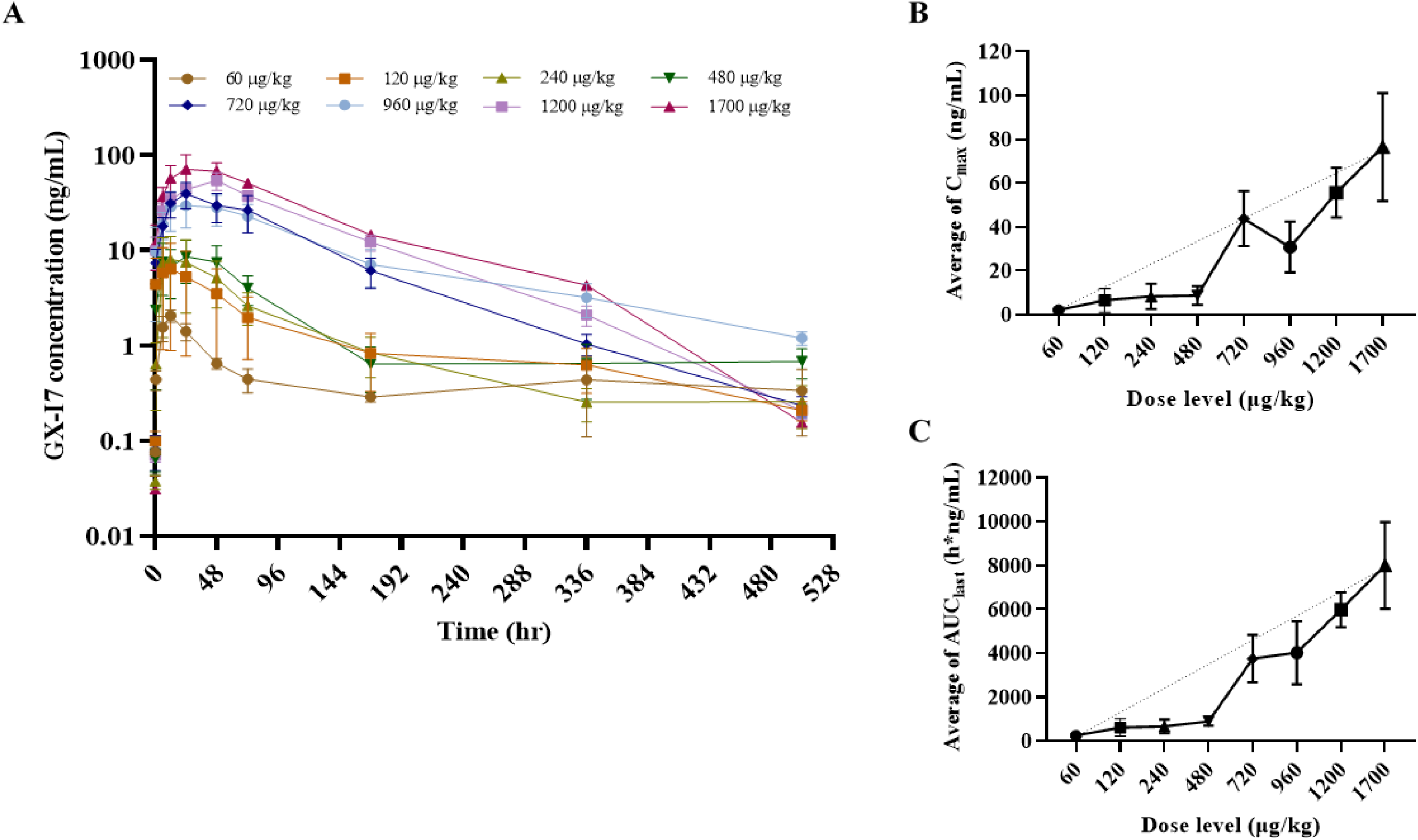
Pharmacokinetics profiles following the first intramuscular administration of GX-I7 ranging from 60 to 1700 μg/kg. (A) The mean serum concentration-time profiles of GX-I7, (B) the maximum concentration (C_max_) versus dose of GX-I7, and (C) the area under the serum concentration-time curve from time zero to the last measurable time-point (AUC_last_) versus dose of GX-I7. the results are presented as mean ± standard deviation (SD).

### Pharmacodynamics (PD)

PD analyses were performed for changes in the absolute lymphocyte count (ALC) and lymphocyte subsets. The ALC, CD4^+^ and CD8^+^ T cell counts showed a statistically significant increase at C1D21 compared to the baseline at all dose levels:60-120, 240-480, 720-960 and 1200-1700 μg/kg groups (*p<0.05, **p<0.01, ***p<0.001, Figure 3A) in a dose-dependent manner. In contrast, GX-I7 administration did not cause significant changes in B or NK cells (Figure 3B). These results are consistent with previously reported findings (23). Increases in ALC were observed in both lymphopenic and non-lymphopenic patients (*p<0.05, ****p<0.0001, respectively; Figure 3C).

**Figure 3.**
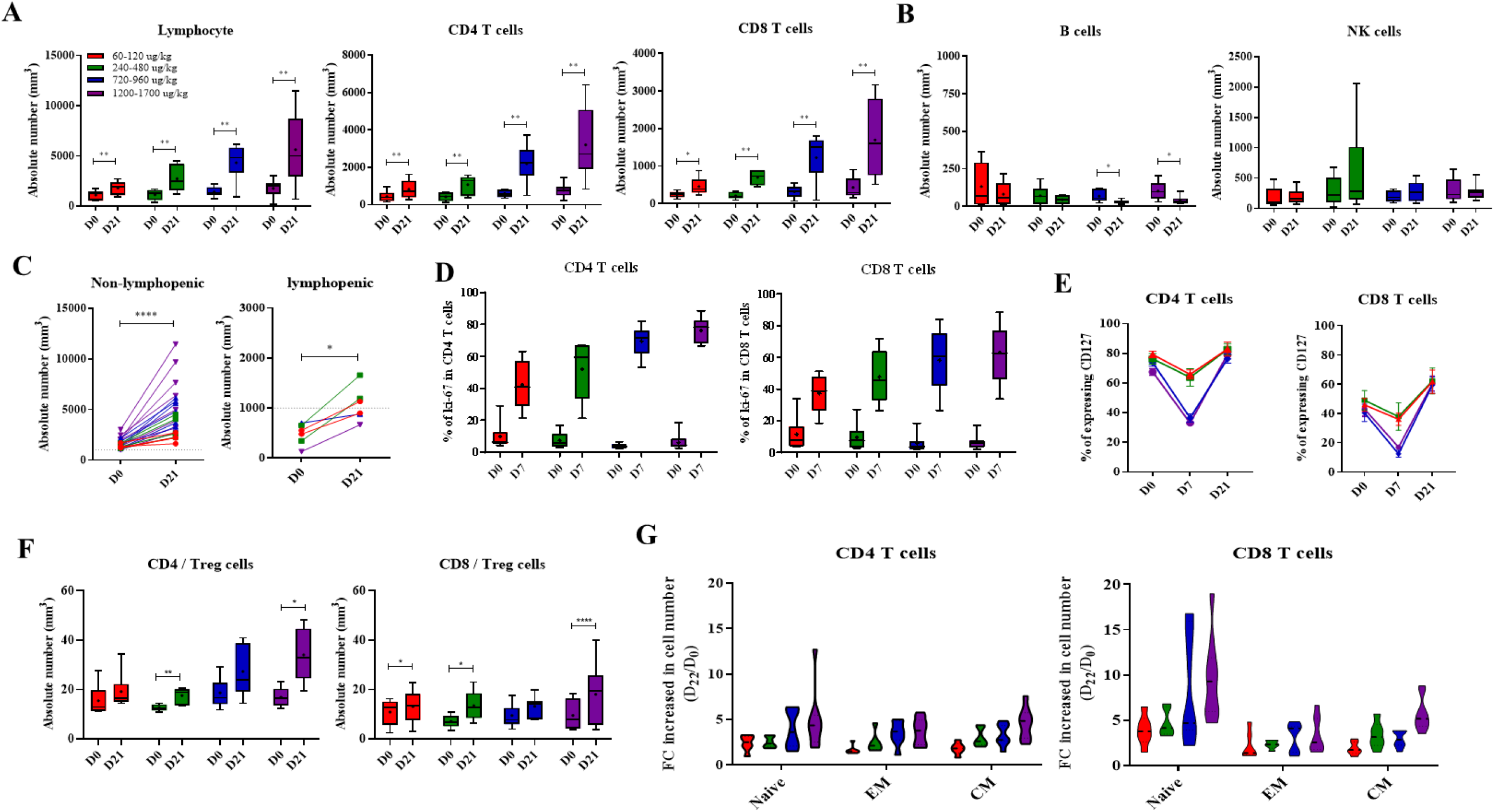
Pharmacodynamic profiles following the first intramuscular administration of GX-I7 across four groups with difference doses. Changes in (A) ALC, CD4^+^, CD8^+^ T cells, (B) other immune cells (C) ALC in non-lymphopenic (ALC>1,000 cells/mm^3^) patients and lymphopenic (ALC<1,000 cells/mm^3^) patients, (D) percentage (%) of Ki67^+^CD4^+^ and Ki67^+^CD8^+^ T cells, (E) percentage (%) of CD127^+^CD4^+^ and CD127^+^CD8^+^ T cells, (F) CD4+T/Treg and CD8^+^T/Treg cells ratios and (G) sub-sets of CD4^+^ and CD8^+^ T cells. *p<0.05, **p<0.01, ***p<0.001 versus baseline (Day 0) group by Wilcoxon matched-pairs signed rank test. FC; fold change.

To evaluate the potential mechanism by which GX-I7 increases lymphocyte counts, Ki67^+^CD4^+^ T cells and Ki67^+^ CD8^+^ T cells were measured. Their percentage peaked on day 7 after treatment and showed a dose-dependent and statistically significant increase in all the groups (Figure 3D). IL-7 binds to IL-7 receptor α (CD127) expressed on the surface of T cells, and CD127 is downregulated or internalized when T cells are over-activated by IL-7 (transcytosis). Indeed, there was a significant but transient decrease in CD127^+^CD4^+^ and CD127^+^ CD8^+^ T cells on day 7 compared with day 0 in each group. However, these decreases were restored at three weeks post dose (Figure 3E). Previous clinical studies of rhIL-7 in HIV-infected or refractory cancer patients revealed that the relative frequency of regulatory T cells (T_reg_) expressing low-level CD127 was reduced due to IL-7-induced expansion of conventional T-cells (24, 25). Consistent with previous studies, the ratio of CD4^+^ and CD8^+^ T cells to regulatory T cells on day 21 after GX-I7 administration compared to baseline showed an increasing trend, and the increase was statistically significant in the 240-480 and 1200-1700 ug/kg groups (Figure 3F). Figure 3G shows the average fold-change of CD4^+^ and CD8^+^ T cell subtypes at day 21 after GX-I7 administration compared to the baseline. While all CD4^+^ and CD8^+^ T cell subsets, including naïve, central memory (CM), and effector memory (EM) T cells, showed significantly increased numbers after GX-I7 administration, notable increases in the number were observed for naïve CD8^+^ and CD4^+^ T cells.

Pre and post-treatment tumor biopsies were collected from five patients who consented to the intervention to evaluate changes in the number of tumor-infiltrating lymphocytes (TILs). Multiplex IHC was used in this study, and all biopsies were collected in the third week after the first injection or the second week after the second injection of GX-I7. A statistically significant increase in CD8^+^/CD4^+^ TILs was observed after GX-I7 treatment (720 μg/kg group, 1 patient; 1200 μg/kg group; four patients) (Figure 4A). Figure 4B shows a representative image of tumor tissue from a patient treated at 1200 μg/kg, demonstrating increased CD4^+^ and CD8^+^ T cell infiltration at week 3 compared to baseline.

**Figure 4.**
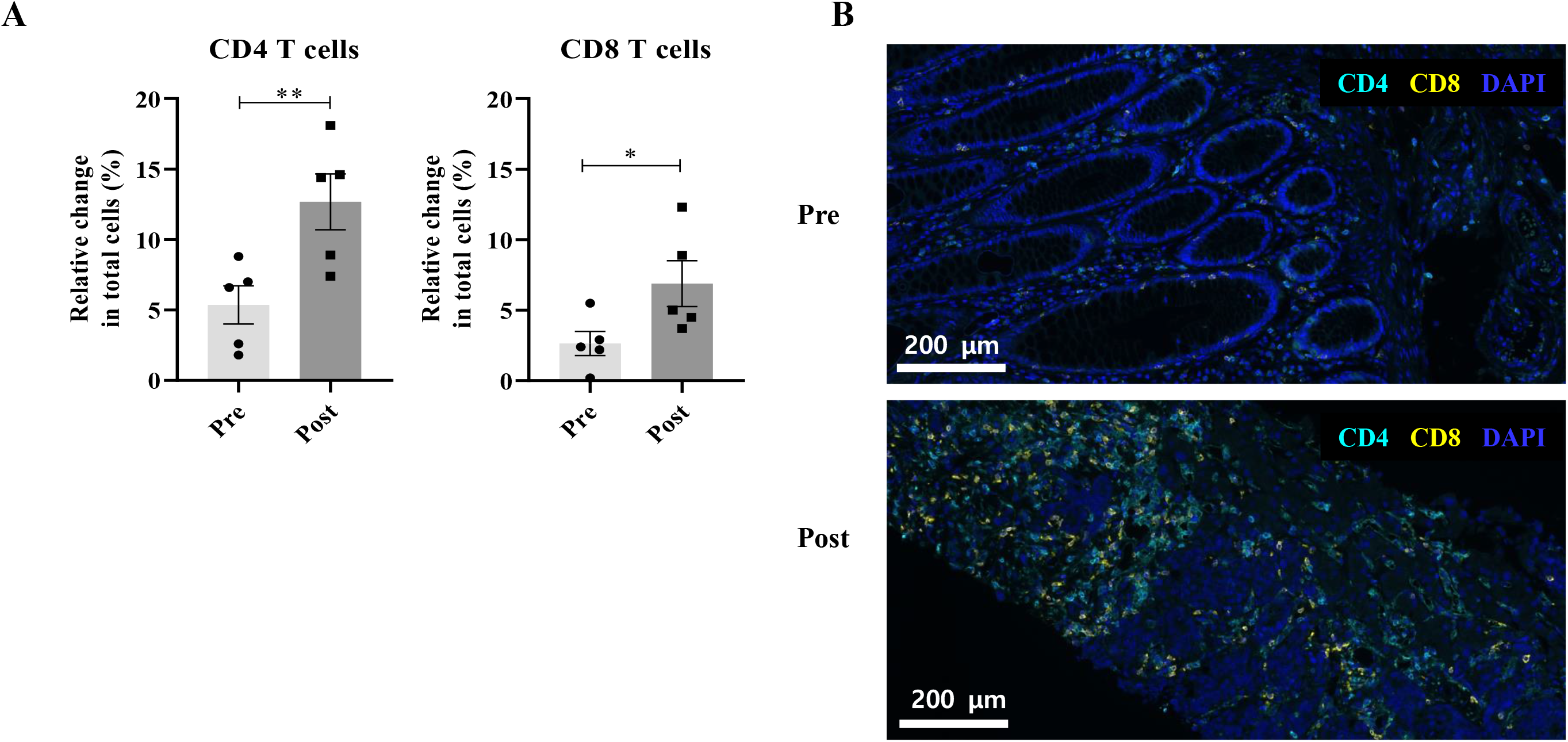
T cell subsets evaluation by Multiplex-IHC for tumor microenvironments at baseline (pre) and GX-I7 treatment (post) from solid tumor patients. All tissue samples were biopsied at the metastasized site, except for one pre-dose sample from one subject. (A) The percentages of infiltrating CD4^+^ and CD8^+^ T cells in the whole tumor samples were higher in post specimens. (B) Representative Multiplex-IHC image of pre-treatment (upper) and post-treatment (lower) in biopsied specimens. Scale bars: 200 μm for IHC and DAPI (blue) was used for nuclear staining. *p<0.05, **p<0.01 versus baseline (Day 0) group by Paired t-test.

## DISCUSSION

This is the first phase 1 study to evaluate the safety, tolerability, pharmacokinetics (PK), and pharmacodynamics (PD) of GX-I7 in patients with locally advanced or metastatic incurable solid tumors. GX-I7 was shown to be safe and well tolerated in a wide range of doses, ranging from 60 to 1200 μg/kg, at 3- or 6-week intervals. Tolerability and safety were confirmed in GX-I7 single subcutaneous and intramuscular dose-escalation tests in healthy subjects (23). This clinical trial was conducted to confirm the safety and tolerability of GX-I7 in patients with locally advanced, recurrent, or metastatic incurable solid tumors. The most common adverse event and adverse drug reactions in all dose groups were injection site reactions, which were also observed following the administration of other cytokine products (26). There were no adverse events, such as a serious injection site reaction, which would result in permanent discontinuation of GX-I7 treatment. MTD and RP2D were confirmed to be 1200 μg/kg based on the dose-limiting toxicity in each GX-I7 dose group. The DLT that occurred was one subject with grade 3 or higher hypersensitivity in the 1700 μg/kg group. Since the safety and tolerability of GX-I7 up to 1200 μg/kg have been established in patients with chemorefractory solid tumors, it is expected that it can be safely administered in combination with other anticancer drugs. For efficacy analysis, anti-tumor activity was assessed in 32 patients who received at least one dose of GX-I7 and completed one cycle of treatment with measurable disease. Six patients (18.75%) showed stable disease (SD), and the median progression-free survival (mPFS) was 5.69 weeks (95% CI:5.13 - 5.98 weeks), with similar mPFS between groups. GX-I7 is a safe and effective T-cell amplifier. Although the primary objective of the study was safety and the number of patients enrolled was low, GX-I7 monotherapy showed a modest clinical benefit, with six patients having stable disease, indicating the need for further studies, in monotherapy or in combination with other immunotherapies, to establish its clinical efficacy in larger trials.

The PK profile provided evidence that GX-I7 is a long-acting IL-7 with a dose-dependent increase in systemic exposure, as indicated by C_max_ and AUC_last_. PD analyses demonstrated that GX-I7 is capable of increasing the absolute lymphocyte counts (ALC), CD4^+^ and CD8^+^ T cells in a dose-dependent manner but does not affect B cell counts. The increase in ALC, as assessed by ΔE_max_ and ΔAUEC, was positively correlated with systemic exposure to GX-I7, as indicated by C_max_ and AUC_last_ (Supplementary Figure S1). In five available paired biopsies, GX-I7 also showed the ability to increase the number of tumor-infiltrating lymphocytes (TILs) of the CD4 and CD8 subtypes. ADA emergence was observed regardless of the dose and dosing interval, but it did not have an impact on the safety of GX-I7. The final RP2D was determined to be 1200 μg/kg based on the maximum observed change in ALC, used as a pharmacodynamic marker, within a dose range that demonstrated safety and tolerability. The increase in Ki67 expression and downregulation of CD127 provided evidence that the increase in ALC and T cells is due to the direct mechanism of action of GX-I7. Administration of GX-I7 resulted in an improvement in the CD4^+^ T cell/T_reg_ and CD8^+^ T cell/T_reg_ ratios, generating a favorable environment for enhanced immune activity.

In this study, most patients received at least two or more doses of GX-I7 at 3-week intervals. However, starting from the second repeated dosing, there was no observed increase in proliferation markers such as Ki67 in T cells and ALC (Supplementary Figure S2A). Based on these results, GX-I7 was administered at 6-week intervals at a dose of 1200 μg/kg in the expansion phase. As shown in Supplementary Figure S2B, the levels of Ki67^+^CD4^+^ and Ki67^+^CD8^+^ T cells, as well as the ALC level (although data from only one patient was available), increased in the blood following repeated dosing of GX-I7 at 6-week intervals. It was not possible to obtain long-term follow-up results of repeated dosing, as most of the patients participating in this clinical trial had late-stage cancer with very short life expectancies. For this reason, an animal study was conducted as a supportive measure in normal cynomolgus monkeys, and it showed that when the monkeys were repeatedly administered GX-I7 at a dose of 3 mg/kg with intervals of 6, 9, and 12-weeks, the proliferation markers and ALC showed a tendency to increase when administered at intervals of 6-week or longer (Supplementary figure S3). Therefore, the dosing interval can be considered an important factor that may affect the PD profile of GX-I7. Subsequent studies with GX-I7 administered GX-I7 with dosing intervals of 6 weeks or longer. ADA occurs in more than 90% of patients after receiving a dose of GX-I7, which is consistent with previous findings in healthy adults. In Phase 1 clinical trials in healthy volunteers, ADA was observed in most patients (75-100%) approximately 10 days after GX-I7 administration, but most of these antibodies disappeared within 330 days of follow-up.

In the present study, repeated administration of GX-I7 at 6-week intervals demonstrated an increase in proliferation markers, despite the presence of ADA. Similarly, in the monkey study, repeated administration of GX-I7 for 6-week or longer resulted in increased expression of proliferation markers, even in the presence of ADA (Supplementary Figure S3, Supplementary Table S3). These results indicate that ADA production does not affect the ability of GX-I7 to increase ALC; however, further studies involving a larger number of patients with longer treatment durations are warranted to confirm these findings. The main pharmacodynamic response of GX-I7 in biological systems is an increase in ALC. However, it is still under investigation whether this GX-I7-related boost in ALC directly leads to increased TIL and mediates clinical efficacy. In the present study, as part of this investigation, multiplex IHC analysis was performed on a small number of patients whose tumors were biopsied, and an increase in CD4^+^ and CD8^+^ T cells was observed within the tumor tissues at 3 or 5 weeks after GX-I7 administration. Thus, GX-I7 as a T-cell amplifier provides a unique opportunity for immuno-oncology combination strategies by reconstituting persistent T-cells.

These findings support the use of GX-I7 as a safe and effective T cell-amplifying agent for correcting T cell deficiency. Studies are ongoing to evaluate the clinical efficacy of GX-I7 when used in combination with other anti-cancer agents. Novel combinations with GX-I7 have the potential to improve the tumor microenvironment and enhance the efficacy of ICI and other anti-cancer treatments to fight cancer.

## Data Availability

All data produced in the present study are available upon reasonable request to the authors
All data produced in the present work are contained in the manuscript

## ACKNOWLEDGMENTS

This work was supported by Bio-Teller, who provided scientific advice for this clinical research.

