## Supplemental Table 1 for "GX-I7(rhIL-7-hyFc, efineptakin alfa), a long-acting IL-7, safely and effectively increased peripheral CD8^+^ and CD4^+^ T cells and TILs in patients with solid tumors"

**Supplementary material**

***Flow cytometry***

Flow cytometry was performed to assess the expression of cell surface and intracellular molecules in cryopreserved PBMCs collected at the specified time points. Surface molecules were stained with antibodies at room temperature (RT) for 15 min. The following anti-human fluorochrome-conjugated antibodies were used: Alexa Fluor 700-conjugated antibody to human CD3 (UCHT1); anti-IL-7Rα-BV421 (A019D5), anti-CD4-PE-Cy5 (OKT4), anti-CD45RA-APC-Cy7 (HI100), anti-Fas-BV650 (DX2), anti-CCR7-BV785 (G043H7), anti-CD31-Alexa Fluor 488 (WM59), anti-TCR γ/δ-Percp-Cy5.5 (B1), anti-CD25-BV650 (BD96; BioLegend), anti-CD19-BV711 (SJ25C1), anti-CD14-BV711 (MφP9), anti-CD3-BV510 (HIT3a), anti-CD8-Alexa Fluor 700 (RPA-T8; all from BD Biosciences); anti-CD19-PE-eFluor610 (HIB19); anti-CD14-PE-eFluor610 (61D3), and anti-CD56-APC (CMSSB; Thermo Fisher). Simultaneously, dead cells were stained using a Live/Dead Fixable Red Dead Cell Stain Kit (Thermo Fisher Scientific). The cells were fixed and permeabilized using a Foxp3 Staining Buffer Set (Thermo Fisher), and intracellular molecules were stained (4 °C, 20 min) using anti-Bcl-2-Alexa Fluor 488 (100; BioLegend), anti-Ki-67-PE-Cy7 (20Raj1), and anti-Foxp3-PE (236A/E7; Thermo Fisher). Acquisition was performed using an LSR II instrument (BD Biosciences) and data analysis was performed using FlowJo software (FlowJo, LLC).

***Multiplex immunohistochemistry***

All biopsies were collected in the third week after the first injection or in the second week after the second injection of GX-I7. Liver, colon, lymph node, and lung biopsy specimens were fixed in formalin, embedded in paraffin (FFPE), and sliced into sections. Slides were heated for at least one hour in a dry oven at 60 °C, followed by multiplex immunofluorescence staining using a Leica Bond Rx™ Automated Stainer (Leica Biosystems, Germany). Briefly, the slides were deparaffinized with Leica Bond Dewax solution (#AR9222, Leica Biosystems), followed by antigen retrieval using Bond Epitope Retrieval (#AR9640, Leica Biosystems). Staining was performed in sequential rounds of blocking with antibody diluent solution (ARD1001EA, Akoya Biosciences, United States), followed by incubation with the primary antibody and OPAL polymer HRP (ARH1001EA, Akoya Biosciences) for 30 and 10 min, respectively. The primary antibodies used were CD4 (ab133616, R&D) and CD8 (MCA1817, Bio-Rad). Nuclei were stained with DAPI (62248, Thermo Scientific, United States) Several regions of interest (ROI) selected by a pathologist, were scanned using the Vectra Polaris Automated Quantitative Pathology Imaging System (Akoya Biosciences) at 20 X magnification. The number of TIL was calculated using Image Analysis software. Analyses were performed on PrismCDX Co., Ltd..

**Supplementary Table S1.** Best Overall Response (BoR) by RECIST v1.1 and Progression Free Survival.

| Group | GX-I7 60 µg/kg (n=3) | GX-I7 120 µg/kg (n=3) | GX-I7 240 µg/kg (n=3) | GX-I7 480 µg/kg (n=3) | GX-I7 720 µg/kg (n=5) | GX-I7 960 µg/kg (n=3) | GX-I7 1200 µg/kg (n=10) | GX-I7 1700 µg/kg (n=2) | Total (n=32) |
|---|---|---|---|---|---|---|---|---|---|
| **BoR, N (%)** | | | | | | | | | |
| Complete Response | 0 | 0 | 0 | 0 | 0 | 0 | 0 | 0 | 0 |
| Partial Response | 0 | 0 | 0 | 0 | 0 | 0 | 0 | 0 | 0 |
| Stable Disease | 1(33.33) | 1(33.33) | 1(33.33) | 0 | 1(20.00) | 0 | 2(20.00) | 0 | 6(18.75) |
| Progressive Disease | 2(66.67) | 2(66.67) | 2(66.67) | 3(100.00) | 4(80.00) | 3(100.00) | 8(80.00) | 2(100.00) | 26(81.25) |
| Not Evaluable | 0 | 0 | 0 | 0 | 0 | 0 | 0 | 0 | 0 |
| **ORR** | | | | | | | | | |
| CR+PR | 0 | 0 | 0 | 0 | 0 | 0 | 0 | 0 | 0 |
| Exact 95% CI | (0.00, 70.76) | (0.00, 70.76) | (0.00, 70.76) | (0.00, 70.76) | (0.00, 52.18) | (0.00, 70.76) | (0.00, 30.85) | (0.00, 84.19) | (0.00, 10.89) |
| **Progression Free Survival (weeks)** | | | | | | | | | |
| Median | 6.26 | 5.69 | 5.69 | 5.13 | 5.55 | 4.13 | 5.84 | 4.91 | 5.69 |
| 95% CI | (5.13, 18.93) | (3.13, 12.39) | (5.27, 17.65) | (1.57, 5.98) | (5.13, 23.92) | (1.99, 6.26) | (2.70, 6.98) | (4.13, 5.69) | (5.13, 5.98) |

**Supplementary Table S2.** PK parameters.

| Dose (μg/kg) | $T_{max}$ (h) | $t_{1/2}$ (h) | $C_{max}$ (ng/mL) | $AUC_{last}$ (h·ng/L) | |
|---|---|---|---|---|---|
| | | | | **Mean** | **Geomean** |
| **60** | 11 (6-11) | 139.7 (80.7-351.3) | 2.11±0.47 | 118.4±135.3 | 214,4±1.69 |
| **120** | 12 (11-72) | 123.2 (104.6-223.6) | 6.53±9.42 | 304.6±701.7 | 386.5±3.12 |
| **240** | 47 (11-48) | 89.2 (76.0-132.9) | 8.30±10.0 | 329.2±550.3 | 511.4±2.42 |
| **480** | 24 (6-24) | 106.4 (44.9-227.3) | 8.66±7.17 | 444.3±350.7 | 836.9±1.55 |
| **720** | 47.5 (12-72) | 78.3 (58.1-121.6) | 43.7±7.77 | 1874.6±2648.3 | 3230.6±1.73 |
| **960** | 24 (11-71) | 87.6 (84.1-91.0) | 30.8±20.1 | 2409.2±2488.5 | 3359.4±2.21 |
| **1200** | 47 (23-72) | 60.8 (43.9-203.3) | 55.6±39.4 | 2994.5±2764.0 | 5350.7±1,69 |
| **1700** | 35.5 (23-48) | 69.7 (53.7-85.7) | 76.5 ±34.8 | 4006.4±2806.1 | 7752.5±1.43 |

**Supplementary Table S3.** Anti-drug antibody (ADA) after GX-I7 administration in normal monkeys.

| **Administration** | **q6w** | | | **q9w** | | | **q12w** | | |
|---|---|---|---|---|---|---|---|---|---|
| Animal # | XN1M01 | XN1M02 | XN1M03 | XN2M01 | XN2M02 | XN2M03 | XN3M01 | XN3M02 | XN3M03 |
| **d-8** | - | - | - | - | - | - | - | - | - |
| **d0** | - | - | - | - | - | - | - | - | - |
| **d7** | - | - | - | - | - | - | - | - | - |
| **d14** | + | + | + | + | + | + | + | + | + |
| **d21** | + | + | + | + | + | + | + | + | + |
| **d28** | + | + | + | + | + | + | + | + | + |
| **d35** | + | + | + | N/A | N/A | N/A | N/A | N/A | N/A |
| **d42** | + | + | + | + | + | + | + | + | + |
| **d49** | + | + | + | N/A | N/A | N/A | N/A | N/A | N/A |
| **d56** | + | + | + | + | + | + | + | + | + |
| **d63** | + | + | + | + | + | + | N/A | N/A | N/A |
| **d69** | + | + | + | + | + | + | N/A | N/A | N/A |
| **d77** | + | + | + | + | + | + | + | + | + |
| **d84** | + | + | + | + | + | + | + | + | + |
| **d91** | + | + | + | + | + | + | + | + | + |
| **d98** | + | + | + | N/A | N/A | N/A | + | + | + |
| **d105** | + | + | + | N/A | N/A | N/A | + | + | + |
| **d112** | + | + | + | N/A | N/A | N/A | + | + | + |

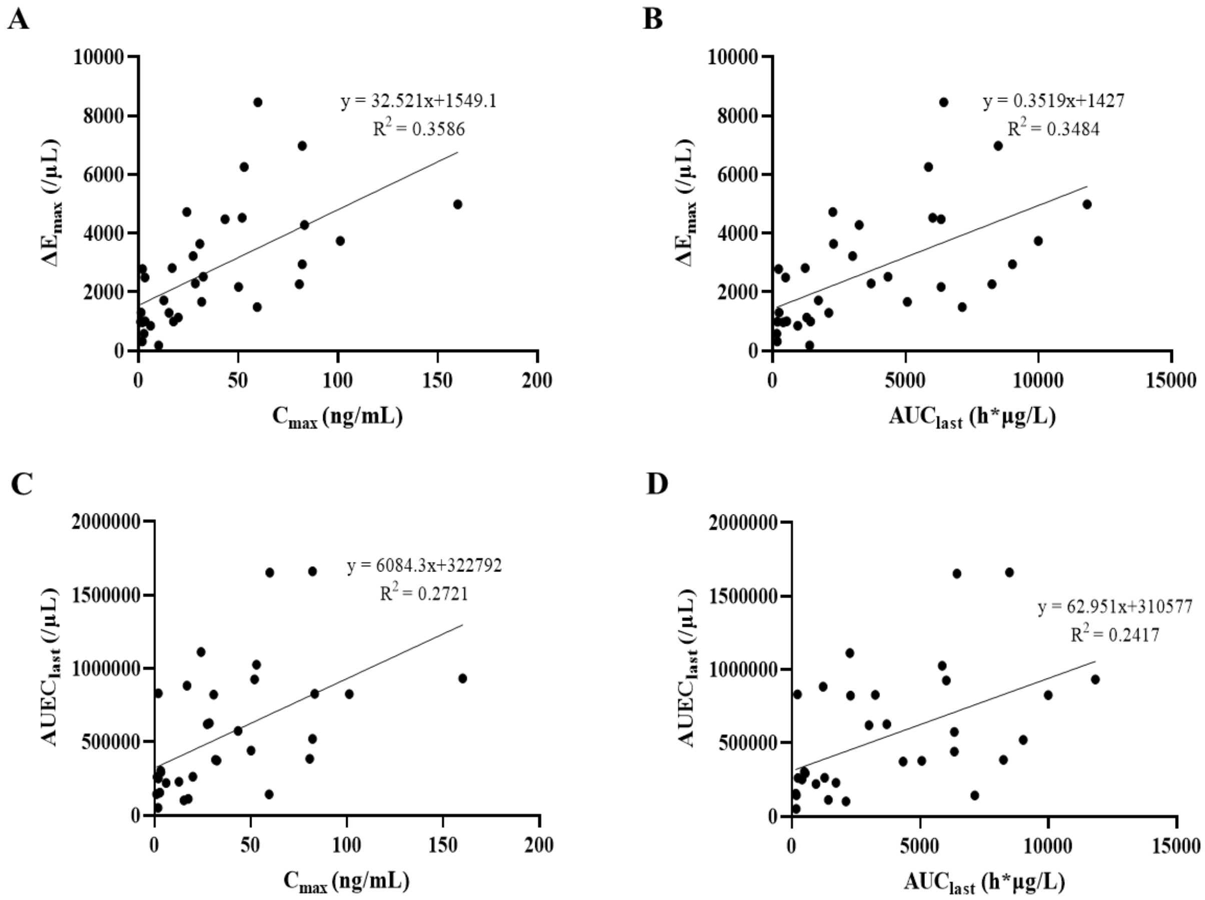

**Supplementary Figure S1.** Relationships between individual pharmacokinetic parameters vs. pharmacodynamic parameters. (A) Maximum concentration (Cmax) of GX-I7 versus maximum absolute lymphocyte count (ΔEmax), (B) Area under the serum concentration-time curve from time zero to the last measurable time-point (AUClast) of GX-I7 versus ΔEmax, (C) Cmax of GX-I7 versus absolute lymphocyte count-time curve until the sampling time at week 3 (ΔAUEC), (D) AUClast of GX-I7 versus ΔAUEC after GX-I7 administration.

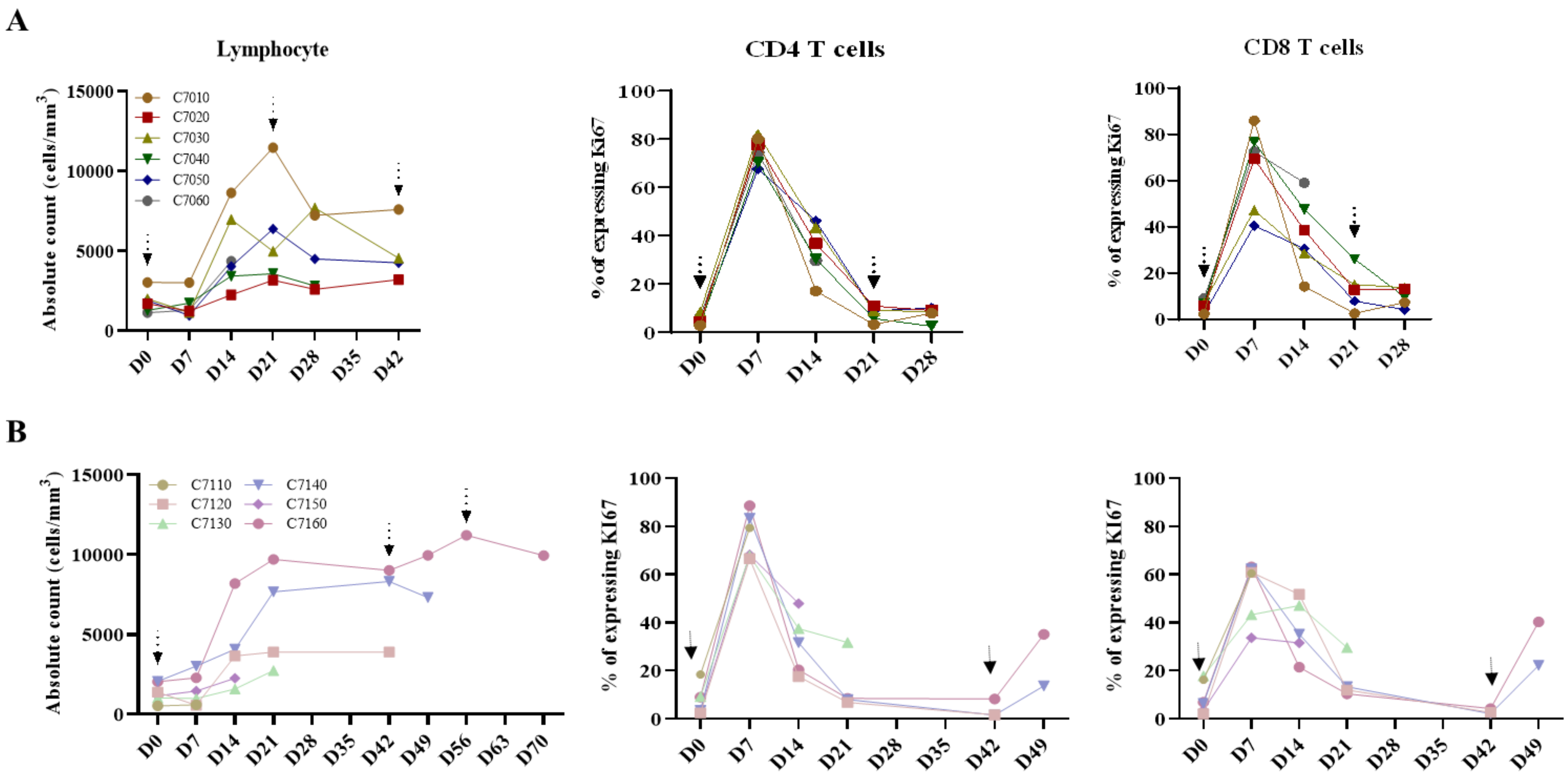

**Supplementary Figure S2.** The changes in ALC, and in ki67 expression on CD4+/CD8+ T cells depending on the GX-I7 dosing interval in patients. (A) GX-I7 administration at 3-week intervals and (B) 6-week intervals. Arrows indicate the time of dosing.

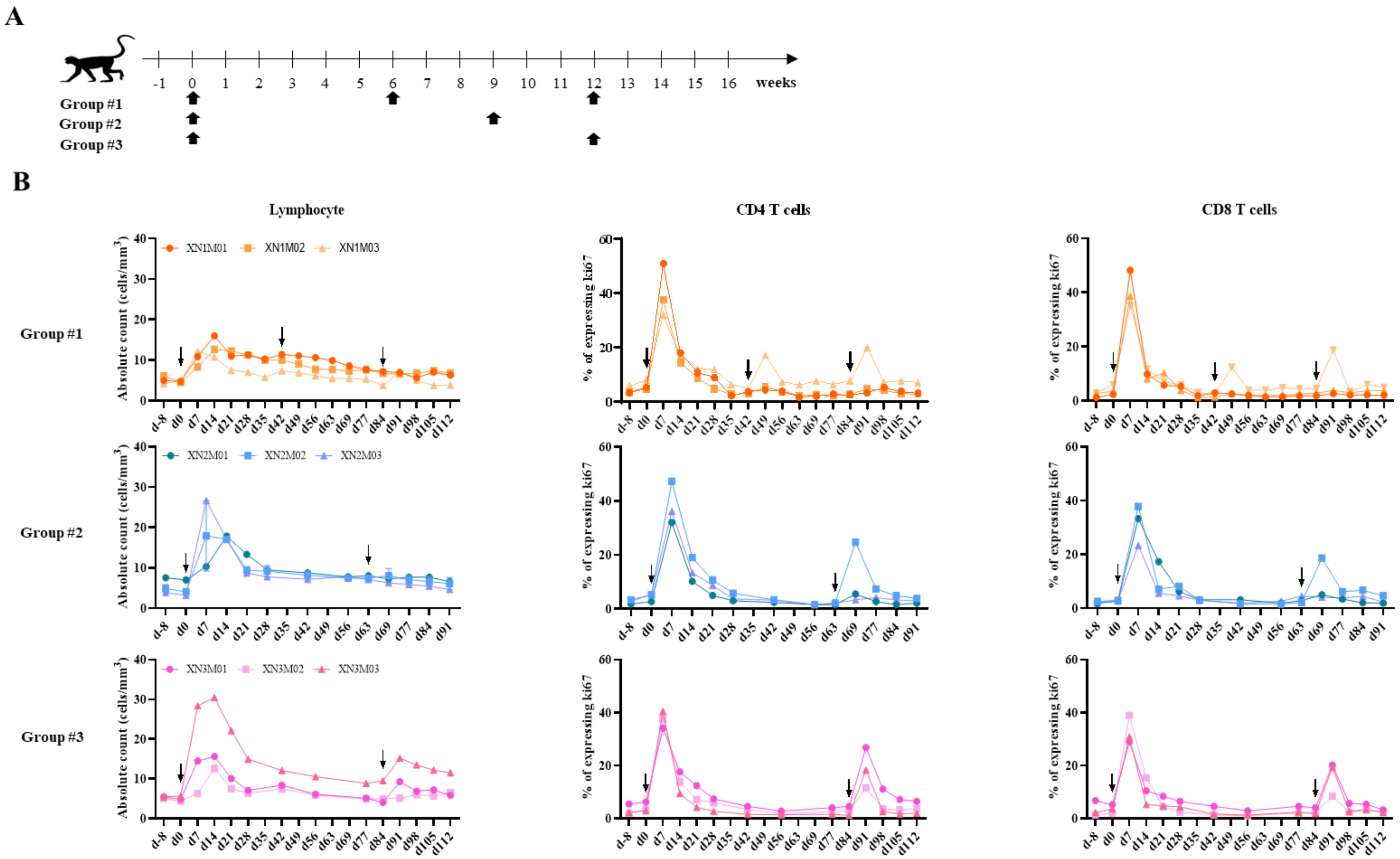

**Supplementary Figure S3.** The changes in ALC, and in ki67 expression on CD4+/CD8+ T cells depending on the GX-I7 injection interval in normal monkeys. (A) Study design, (B) GX-I7 injection at 6-week intervals (upper), 9-week intervals (middle), and 12-week intervals (lower). Arrows indicate the time of dosing.
